# Concerns with the use of imputation to assign HLA allele-level typing in research predicting transplant outcomes

**DOI:** 10.1101/2020.07.19.20157461

**Authors:** Anat R Tambur, Michael Gmeiner, Charles F Manski

**Affiliations:** Feinberg School of Medicine, Northwestern University, Chicago, IL 60611; Department of Economics, Northwestern University, Evanston, IL 60208; Institute for Policy Research, Northwestern University, Evanston, IL 60208

## Abstract

Incomplete information on HLA allele typing is a persistent problem when analyzing the role of Human Leukocyte Antigen (HLA) in transplantation. To refine the predictions possible with partial knowledge of HLA typing, some researchers use HaploStats statistics on the frequencies of haplotypes within specified ethnic/national populations to impute complete HLA allele typing. We evaluated methods that use imputation to predict patient outcomes after organ transplantation, with focus on prediction of graft survival conditional on typing information of the donor and recipient. Logical arguments show that imputation yields no predictive power when predictions are conditioned on all observed HLA typing data. Computational experiments indicate that imputation does not have predictive power when applied to risk-assessment models that make predictions conditional on only part of the observable HLA data. We therefore caution against reliance on imputation to overcome incomplete measurement. We encourage high-resolution typing of HLA antigens to improve prediction of transplant outcomes and matching of donors with recipients. Similar considerations should likely apply in other clinical settings.

## 1. Introduction

The Final Rule requires the establishment of policies for organ allocation that will “seek to achieve the best use of the donated organs”^1^. As such, an important problem is to predict the outcomes that occur when an organ is transplanted into a recipient. Observed predictors include data characterizing the organ and patient. A prominent consideration is the genetic match between donor and recipient, as measured by their Human Leukocyte Antigen (HLA) genotypes. Research in transplant immunology has shown that, all else equal, probability of graft survival declines with the degree of HLA mismatch between the donor and recipient.^2 3 4^

A persistent problem in clinical practice as well as in research analyzing historical data in aggregate is incomplete information on donor and recipient HLA typing (no data on some loci and lack of high-resolution typing). The Organ Procurement and Transplantation Network (OPTN) requires transplant centers to provide pre-transplant data to the Scientific Registry of Transplant Recipients (SRTR), which collates the data with reports of transplant outcomes and makes the combined data available for analysis. Researchers use the SRTR data to investigate how transplant outcomes vary with measured attributes of organ quality, patient age/health, and HLA typing.

Until 2010, the only HLA typing information collected for donors and recipients was on HLA (A, B, DR) loci, at the serologic level. With initiation of cPRA measurement, molecular typing was required. Over time, additional loci information was mandated for the donor – including HLA (DQB1, DPB1, DQA1). Importantly, no such requirements were made for the typing of patients. Many patients are listed for transplant with only HLA (A, B, DR) low-resolution typing information available, although some patients have serologic equivalent DQ typing.

The technology to determine HLA typing in a high-resolution/allele-level HLA (A, B, C, DRB1, DRB3/4/5, DQA1/DQB1, DPA1/DPB1) typing exists, but not all typing ambiguities can be resolved within the time constraints imposed by deceased donor organ allocation. Higher resolution typing would be beneficial because each low-resolution, two-digit, typing represents multiple alleles, whereas each four-digit typing identifies a unique allele. Donor and recipient HLA types that appear matched with two-digit coding may be mismatched with four-digit coding. A patient may have antibodies to an allele within a low-resolution antigen group, but the donor typing may be of a different allele, against which the recipient does not have a donor-specific antibody (DSA). Differences at the allele level can also translate into differences in the assignment of molecular mismatches, currently proposed for use in risk stratification of transplant recipients.

Aiming to refine the predictions possible with incomplete knowledge of HLA data, methods have been developed to use available statistics on the distributions of high-resolution HLA typing within specified ethnic/national sub-populations to impute unobserved typing. Two prominent approaches are imputation of most prevalent types (“winner-take-all”) and random multiple imputation (RMI). Both approaches use the distribution of high-resolution typing given low-resolution typing.

The most comprehensive database providing frequency distributions of HLA alleles and haplotypes is HaploStats (http://www.HaploStats.org).^5^ HaploStats facilitates access to HLA genotype frequency data for various ethnic/national groups. A researcher or clinician can input the available, incomplete, typing data for a donor or recipient who is a member of a specified group. HaploStats accesses its genotype frequency data and outputs the frequency distribution of high-resolution typing (HLA-A, B, C, DRB1, DQB1) conditional on the data provided. The frequency distribution output by HaploStats is formatted in order of the prevalence of specific haplotypes.

Given that much of the transplant related data available within the OPTN and the SRTR is associated with low-resolution typing data only, and retrospective typing of donor-recipient pairs is impractical, using HaploStats to impute high-resolution typing may seem an appealing way to overcome incompleteness of observed HLA data. However, the appeal does not survive under scrutiny. In this paper, we caution against reliance on imputation to cope with incomplete measurement of HLA types.

Sections 2 and 3 show that, in principle, imputation cannot improve prediction beyond the information provided by observable HLA antigens. Haplostats provides frequency distributions of four-digit HLA types conditional on the observed two-digit HLA data that researchers input. Imputed four-digit types, whether computed in the winner-take-all manner or by drawing at random from Haplostats distributions, vary systematically only with the observed two-digit data that researchers input. They logically cannot vary systematically with patient outcomes. Formally, imputations are statistically independent of patient outcomes, conditional on the observed two-digit HLA data that researchers use to produce them. Hence, imputations cannot improve prediction of outcomes beyond the information in observable HLA data.

If researchers were able to make statistically precise predictions of transplant outcomes conditional on all observable HLA data, the above logical argument would suffice to conclude that the use of imputation should be abandoned. In practice, we cannot draw such a stark conclusion based on logic alone. The reason is that the finite number of transplant cases observed in the SRTR files makes it impractical to predict patient outcomes conditional on all observable HLA data. Hence, researchers perform substantial dimension reduction when predicting transplant outcomes.

The number of distinct two-digit (donor, recipient) HLA types is large. Hence, the number of SRTR cases is small for most types. As a result, attempting to predict outcomes conditional on all observable HLA data yields highly imprecise findings for most two-digit types. Recognizing this, researchers predicting transplant outcomes uses risk-assessment models that make predictions conditional on only a small part of the observable (donor, recipient) HLA data.

In particular, it is common to predict graft survival conditional on the number of (donor, recipient) HLA mismatches at various loci, rather than on the underlying HLA types. Many combinations of observed two-digit types can yield the same number of two-digit mismatches. Hence, conditioning only on the number of mismatches improves the statistical imprecision of estimated models, albeit at the cost of not using all available HLA information.

When researchers perform risk assessment with models that condition only on the number of mismatches, it is theoretically possible that imputations may have predictive power. The frequency distributions of four-digit types generated by HaploStats condition on the two-digit HLA data that researchers input, not only on the number of mismatches implied by these data. Hence, imputations might possibly add predictive power to that attainable conditioning only on number of mismatches.

It does not seem possible to determine theoretically the predictive power that imputations may have when used as inputs to various risk-assessment models. Given this, we have performed computational experiments that illustrate what happens in some realistic scenarios. These experiments, reported in Section 4, suggest that imputation does not yield predictive power in practice.

## 2. Winner-Take-All Imputation of Most Prevalent Types

Literature search demonstrates that most publications using imputation to address missing HLA data chose the most prevalent type approach.^6 7 8 9^ However, recent studies have raised concerns.^10 11 12^ Even when the most prevalent haplotype accounts for a large portion of the relevant population, the donor or recipient may not actually have the imputed haplotype. The problem is more consequential when less than half the population have the most prevalent haplotype.^13^

When conditioning on all observable donor and patient antigens, the most prevalent types of donors and patients are constant, and therefore have no predictive power over survival. Imputation of most prevalent types could possibly have predictive power when imputing conditional on less stringent conditions. We present empirical results regarding imputation of most prevalent types in Section 4, where we consider less stringent conditions than conditioning on all observable antigens.

## 3. Random Multiple Imputation

In RMI, a researcher replaces the low-resolution HLA data in each SRTR case with a high-resolution value drawn from the appropriate conditional distribution available in HaploStats. These imputations are treated as data, allowing for a predictive transplant-outcome model to be estimated. Due to the uncertainty of the accuracy of a single draw, the process is repeated multiple times, creating multiple pseudo datasets. This results in a distribution of estimates.

RMI has been promoted as a general approach for coping with data whose values are missing at random.^14 15^ It has been used to impute haplotypes in a study predicting onset of chronic lymphocytic leukemia,^16^ one predicting outcomes of kidney transplants,^17^ and was recommended as an approach to handle missing outcome data in transplant trials.^18^

Justifications for some applications of RMI have been developed, drawing on elements of statistical theory. However, justification for application to study the role of HLA in transplantation has been absent. Consider the objective of predicting patient outcomes conditional on attributes characterizing the donor and recipient. A common outcome of interest is the future date of graft failure. Let the outcome of interest be denoted y. The observed attributes characterizing the donor and recipient commonly include demographic characteristics including ethnicity/nationality, a measure of organ quality such as the Kidney Donor Profile Index (KDPI)^19^, and HLA typing obtained from SRTR (A, B, DR, and potentially C and DQ). Let this vector of attributes be denoted x. Let the unobserved high-resolution donor and recipient HLA typing be denoted w.

The researcher would like to learn the distribution of outcomes conditional on the observed attributes and the unobserved complete HLA typing. Let this be denoted P(y|x, w). The SRTR files enable one to learn the distribution of y conditional on x; that is, P(y|x). HaploStats enables one to learn the distribution of w conditional on x; that is, P(w|x).

Let ω denote a random draw from P(w|x). Given a sample of size N, a pseudo sample has the form (yi, xi, ωi), i = 1, …, N. This combines the observed data on (y, x) with the imputed data for the unobserved values of w. One then uses the values of (x, ω) to predict the values of y. For example, a researcher may estimate a regression model. Whatever methodology one uses, one estimates the distribution P(y|x, ω). Multiple imputation means repeating this process of drawing *ω*multiple times.

Our concern is that the estimated predictive distribution P(y|x, ω) does not equal the distribution of interest, P(y|x, w). By construction, the randomly drawn values ω systematically vary only with the observed attributes × of sample members. Conditional on x, realizations of ω are statistically independent of the outcomes y of sample members.

To see this, recall that *ω*is drawn from *P*(*w*|*x*). Therefore, *P*(*y*|*x,ω*) = *P*(*y*|*x,P*(*w*|*x*)). Conditioning on × inherently conditions on everything determined by x, including P(w|x), therefore *P*(*y*|*x,P*(*w*|*x*)) equals P(y|x). The conclusion is that *P*(*y*|*x,ω*) equals P(y|x), the distribution of outcomes directly observed in the data. Thus, random imputation has no predictive power for outcomes beyond what one can achieve by using × alone to predict y.

### 3.1. The Logic of Prediction with Incomplete HLA Data

Given the above, it is natural to ask what predictions of transplant outcomes can logically be made by combining SRTR and HaploStats data. Formally, what can be learned about P(y|x, w) given knowledge of P(y|x) and P(w|x)? This question has been addressed in previous work.^10^ The question has been answered in generality in research on the *ecological inference* problem.

Analysis is simplest when the outcome of interest can take two values, say 0 and 1. For example, y = 1 may denote that a graft survives for a specified length of time and y = 0 that it does not. Then simple application of basic probability theory shows that, for any values (x, w) of the observed and unobserved attributes, the outcome probability P(y = 1|x, w) lies between certain lower and upper bounds that are computable given the available information.

The lower bound is [P(y = 1|x) – P(not w|x)]/P(w|x) and the upper bound is P(y = 1|x)/P(w|x). The lower bound is informative, in the sense of being larger than zero, if P(y = 1|x) exceeds P(not w|x). The upper bound is informative, in the sense of being smaller than one, if P(y = 1|x) is less than P(w|x).

For example, suppose the SRTR data show that when a donor and recipient have observed attributes x, the frequency with which a graft survives for a given length of time is P(y = 1|x) = 0.6. Drawing on HaploStats, suppose that w is the most prevalent pair of haplotypes when the donor and recipient have attributes x, with P(w|x) = 0.8. Then the computable lower bound on P(y = 1|x, w) is (0.6 – 0.2)/0.8 = 0.5 and the upper bound is 0.6/0.8 = 0.75.

## 4. Using Imputation in Risk-Assessment Models that Condition on Number of HLA Mismatches

Conditioning on all observed HLA data, *x*, is extremely difficult due to sample size limitations. Even in the full SRTR dataset, there were few cases with sufficiently many observations to make analysis feasible. Full conditioning being infeasible, researchers use dimension-reduction methods to predict patient outcomes.

A common practice is to condition on the total number of mismatches at the low-resolution level, *m*_2_. The variable *m*_2_is a one-dimensional function of *x*. A researcher may use HaploStats to impute antigens *ω*and then estimate *P*(*y*|*m*_2_,*ω*). In principle, the conditional probabilities *P*(*y*|*m*_2_,*ω*) may have predictive power relative to the probabilities *P*(*y*|*m*_2_), which condition only on the number of mismatches. It is difficult to determine theoretically the extent of predictive power, if any. We can, however, provide computational evidence.

To investigate the potential added value of imputation in risk assessment models we performed computational exercises in an idealized setting in which we know “true” high-resolution HLA typing and “true” survival. The rationale for creating an idealized setting is to provide a known benchmark against which we can evaluate the performance of alternative methods that use imputations in different ways. We use Haplostats to generate synthetic high-resolution HLA typing data, conditional on observed low-resolution data. We specify a model to generate synthetic five-year graft survival data.

We restrict attention to transplants in 2012-2013, these being recent cases for which graft survival can be observed for at least five years. We perform separate analyses for three categories of patient race/ethnicity: White, Black, and Hispanic. We examine 1,600 consecutive transplants of each race/ethnicity. Of these, HaploStats provides haplotype distributions for patients and donors of 1,496 transplants with white patients, 1,370 with black patients, and 1,416 with Hispanic patients. We perform imputation using distributions of high-resolution antigens that condition on two levels of observed low-resolution HLA data: (1) A, B, DR and (2) A, B, C, DR, DQ antigens. The SRTR dataset provides low-resolution data for all five loci.

We create 48 categories of data based on the race/ethnicity of the patient and quartiles of the patient’s age and KDPI within each race/ethnicity group. We calculate the fraction, Z, of individuals in each category whose grafts are observed to survive for at least five years. We treat this as the category’s baseline five-year survival rate. Using the observable, low-resolution antigens of patients and donors, we draw synthetic high-resolution information for patients and donors from their HaploStats distributions. These synthetic data are randomized imputations. However, once drawn, we hold them fixed and view them as “true” antigens in the idealized setting, rather than repeatedly drawing them as in RMI. This explains our use of the term *synthetic* antigens here rather than imputed antigens.

To predict survival, we assign a matching score to each transplant to measure the risk associated with genetic mismatches. We let matching at DQ and DR be worth 1 point for each match, matching at A and B be worth .5 points for each match, and matching at C be worth .25 points for each match. We assume that the probability of graft survival ranges from [Z – 0.1, Z + 0.05]. We linearly assign probabilities in this range such that the transplant with the highest score is assigned survival probability Z + 0.05 and the transplant with the lowest score is assigned probability Z – 0.1. Using the assigned probability of survival, we randomly assign a synthetic binary survival outcome to each patient. This is considered the “true” survival outcome in the idealized setting.

As a baseline, we use maximum likelihood to estimate logit models predicting five-year survival as a function of the observed number of low-resolution mismatches, patient age, and KDPI. This allows us to compare the predictive power of each method using imputations with the predictive power of observable antigens alone.

The right panel of Table 1 reports findings with the synthetic outcome. The estimated coefficients show that the number of mismatches strongly affects survival probabilities and is statistically significant by conventional criteria. These findings should not be surprising. After all, synthetic survival outcomes were generated in a manner that make the idealized survival probability decrease with the number of mismatches.

**Table 1:** Logit Coefficients: Observed Low Resolution Mismatches and Five-Year Survival

|  | Observed Five-Year Survival |  |  | Synthetic Five-Year Survival |  |  |
| --- | --- | --- | --- | --- | --- | --- |
|  | White | Black | Hispanic | White | Black | Hispanic |
| Mismatches 0-10 | 0.0010<br>(0.0261) | 0.0193<br>(0.0341) | -0.0530<br>(0.0329) | -0.0820<br>(0.0255) | -0.1109<br>(0.0341) | -0.1215<br>(0.0308) |
| KDPI | -0.0150<br>(0.0025) | -0.0109<br>(0.0025) | -0.0119<br>(0.0028) | -0.0118<br>(0.0023) | -0.0129<br>(0.0024) | -0.0069<br>(0.0026) |
| Age | -0.0171<br>(0.0050) | 0.0047<br>(0.0048) | -0.0069<br>(0.0049) | -0.0123<br>(0.0048) | 0.0031<br>(0.0048) | -0.0109<br>(0.0047) |
| Constant | 2.8677<br>(0.3235) | 1.3284<br>(0.3423) | 2.7329<br>(0.3192) | 2.8396<br>(0.3028) | 2.3983<br>(0.3437) | 3.0092<br>(0.3245) |
| Observations | 1,496 | 1,370 | 1,416 | 1,496 | 1,370 | 1,416 |

Robust standard errors in parentheses.

For comparison, the left panel reports findings with actual rather than synthetic survival outcomes. Here the estimated coefficients show negligible and statistically insignificant variation in survival probability with the number of mismatches. Thus, within this subset of the SRTR dataset, low-resolution mismatch does not predict survival in the real world.

To evaluate RMI, we estimate logistic regressions of five-year graft survival on imputed high-resolution mismatches at the (A, B, C, DR, DQ) antigens, *ω*, KDPI, and patient age. We draw 200 pseudo samples, each imputing *ω*conditional on low-resolution (A, B, DR). Supplementary material reports the average logit coefficients and the average standard errors across the 200 pseudo samples. It also shows results imputing based on low-resolution (A, B, C, DR, DQ).

We evaluate imputation of most prevalent types by imputing the most common high-resolution antigens for each donor and patient and treating these as data, then estimating logistic regressions that include age and KDPI. Supplementary material shows coefficients when imputation of most prevalent types is performed conditioning on low-resolution (A, B, DR) antigens. It also shows results imputing based on low-resolution (A, B, C, DR, DQ).

To compare the predictive power of each method, we use the logit coefficients to estimate the effect of mismatches increasing by 1 (either real or imputed). We do this for a patient with 0 mismatches, age of 50, and KDPI of 50, and calculate the estimated change in survival probability of increasing mismatches to 1. Results are in Table 2. For RMI, we report the average effect and the average standard error.

**Table 2:**
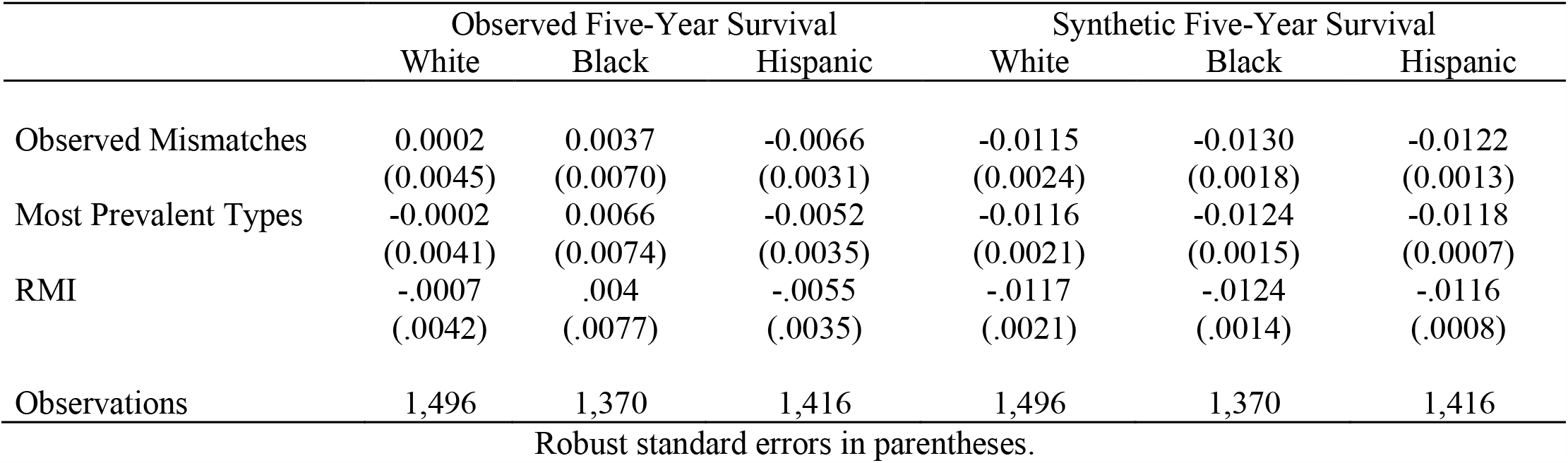
Estimated Effect of an Increase in Mismatches on Survival Probability

|  | Observed Five-Year Survival |  |  | Synthetic Five-Year Survival |  |  |
| --- | --- | --- | --- | --- | --- | --- |
|  | White | Black | Hispanic | White | Black | Hispanic |
| Observed Mismatches | 0.0002<br>(0.0045) | 0.0037<br>(0.0070) | -0.0066<br>(0.0031) | -0.0115<br>(0.0024) | -0.0130<br>(0.0018) | -0.0122<br>(0.0013) |
| Most Prevalent Types | -0.0002<br>(0.0041) | 0.0066<br>(0.0074) | -0.0052<br>(0.0035) | -0.0116<br>(0.0021) | -0.0124<br>(0.0015) | -0.0118<br>(0.0007) |
| RMI | -.0007<br>(.0042) | .004<br>(.0077) | -.0055<br>(.0035) | -.0117<br>(.0021) | -.0124<br>(.0014) | -.0116<br>(.0008) |
| Observations | 1,496 | 1,370 | 1,416 | 1,496 | 1,370 | 1,416 |

Robust standard errors in parentheses.

Figure 1 shows the estimated effects of an additional mismatch for each baseline number of mismatches from 0 to 9. Results are shown for a white patient with age 50 and receiving a kidney with KDPI 50. The x-axis is the number of original mismatches. The y-axis is the estimated effect of an additional mismatch.

**Figure 1.**
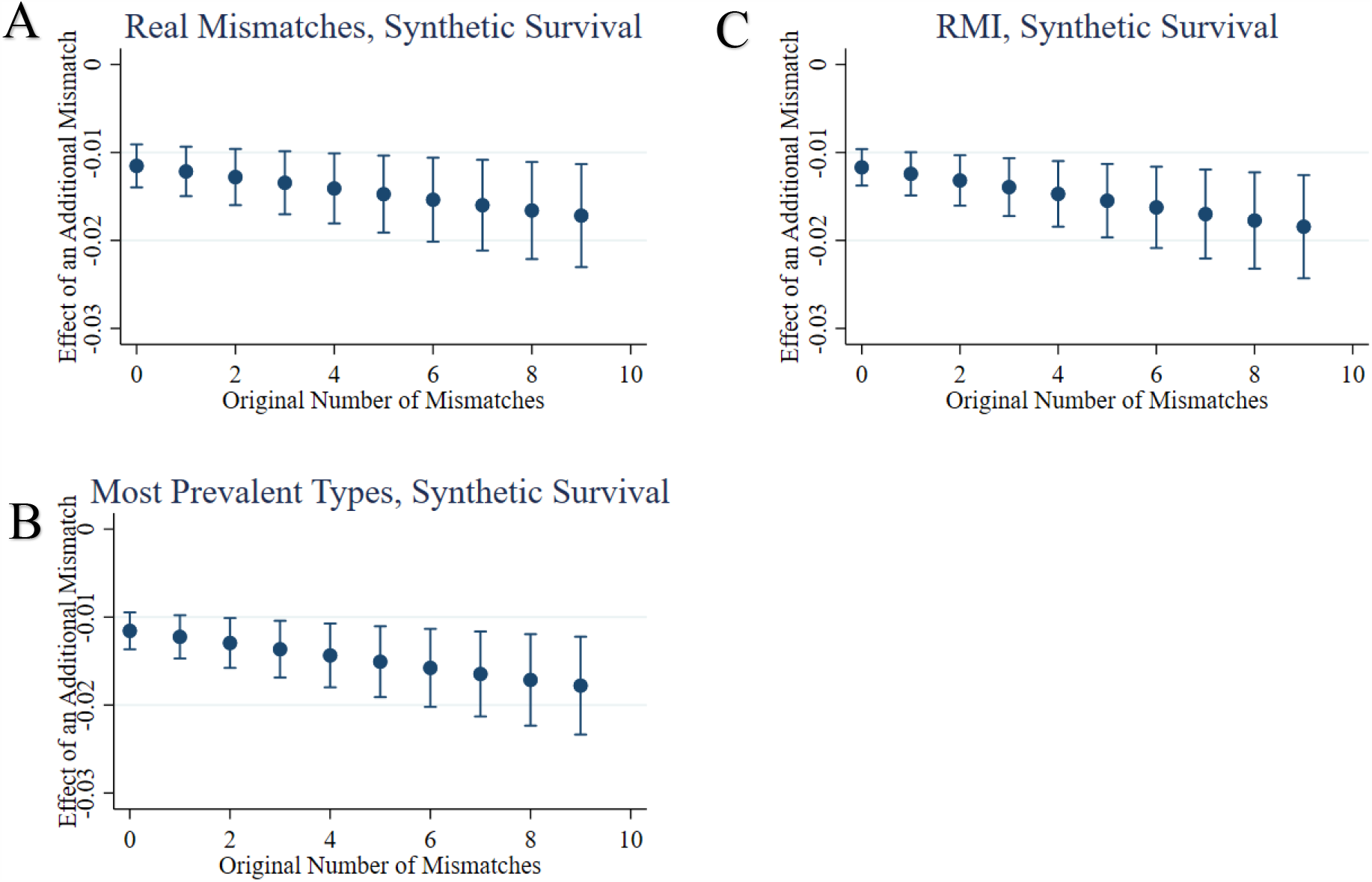
Investigating the potential added value of imputation in risk assessment models: Comparison between real and imputed data. For this exercise we chose a White subject, age of 50, KDPI of 50, zero mismatch with their donor, and computed the effect of adding individual mismatches on their five-year graft survival, presented with standard error. For reference we used synthetic survival by assigning “true” high-resolution antigens given low-resolution antigens, then creating a probability of survival given the synthetic high-resolution mismatches. Panel A provides calculations using coefficients from a logit regression of five-year survival on *observed, low-resolution mismatches*, age, and KDPI. Panel B provides calculation using coefficients from a logit regression of five-year survival on the imputed number of high-resolution mismatches using imputation of *most prevalent types*, age, and KDPI. Panel C provides calculation for the *RMI* approach - For each patient and donor, high-resolution antigens were imputed given low-resolution antigens 200 times. With each draw, a logistic regression was estimated of five-year survival on the number of imputed mismatches, age, and KDPI. Using the average coefficients and standard errors of these 200 regressions, estimates were calculated.

Table 2 and figure 1 show that the predictive power is similar for each method. RMI, imputation of most prevalent types, and using observable low-resolution mismatches, all resulted in similar estimated effects. This suggests that imputation of either form provides no additional predictive power compared to observable data.

## 5. Discussion

We have reported two main findings. When conditioning fully on the observed HLA data, RMI provides no additional predictive power and imputation of most prevalent types is a constant, providing no predictive power. When conditioning only on number of mismatches, RMI and imputation of most prevalent types may have predictive power in principle but do not in practice.

Given these conclusions, it is natural to ask what predictions of transplant outcomes can logically be made by combining SRTR and HaploStats data. This question has been addressed in earlier work,^10^ deriving mathematical bounds on survival probabilities.

One would naturally like to refine the predictions of transplant outcomes currently possible with SRTR and HaploStats data. It has been proposed to enrich information from the SRTR with HaploStats-imputed HLA data to refine simulation of allocation algorithms. RMI and imputation of most prevalent types have been proposed. However, the intuitive appeal of imputation does not hold up to scrutiny. Study of the logic of the prediction problem shows that combining SRTR and HaploStats data may yield informative bounds on outcome probabilities, but it cannot yield precise probabilistic predictions.

While analysis of the prediction problem enables one to make the most of the available data, it would be better to collect more refined and comprehensive HLA data. The reason why the existing SRTR files do not provide complete HLA typing for organ donors and recipients is that the OPTN has not required transplant centers to report this information. This is partly due to the natural evolution of typing methods and partly due to the time it takes to obtain high resolution typing.

HLA laboratories can produce high-resolution typing in many cases, even with current SSO typing techniques. These reports are currently uploaded to UNet/DonorNet and can be viewed for histocompatibility consultation given by HLA lab directors in real time. However, these data are not available for broader analysis as they are not stored by the SRTR. We hope that in the future centers will be required to enter high resolution typing in cases where it is feasible to obtain them, including living donor-recipient pairs and patients on the deceased donor wait list.

Table 1 shows coefficients and standard errors from logit regressions. Each column shows a separate regression using transplants for which the patient’s race is that of the column heading. The left three columns use observed five-year survival as the outcome variable. The right three columns use synthetic survival created by probabilistically assigning high-resolution antigens given low-resolution antigens, then creating a probability of survival given the synthetic high-resolution mismatches. All logit regressions use KDPI, age of the patient and the observed number of low-resolution mismatches as covariates

Table 2 shows the estimated effects, and standard errors, for increasing the number of low-resolution mismatches from 0 to 1 for an individual with KDPI of 50 and age of 50. These effects are calculated using coefficients from logit regressions of five-year survival on age, KDPI, and the number of mismatches. Mismatches are calculated using the method shown in the row title, first being observed low-resolution mismatches, second being imputed high-resolution mismatches using the imputation of most prevalent types, and third being imputed high-resolution mismatches from RMI. The third row is calculated by performing 200 repetitions of RMI logit regressions and averaging coefficients. Each cell shows results from separate estimation using transplants for which the patient is of the race in the column heading. The left three columns use observed survival as the outcome variable. The right three columns use synthetic survival created by probabilistically assigning high-resolution antigens given low-resolution antigens, then creating a probability of survival given the synthetic high-resolution mismatches.

## Data Availability

The standard analysis files from the SRTR are restricted. Haplostats data is freely available using their online tool. The authors can provide code that was used to perform analysis and scrape Haplostats. To fully replicate results of this paper, researchers need to obtain standard analysis files directly from the SRTR.

https://www.srtr.org/

https://www.haplostats.org/

## Supplementary Material

### Supplementary Material 1 – Estimation Results

In section 4 we describe computational results that evaluate RMI by estimating logistic regressions of five-year graft survival, *y*, on imputed high-resolution mismatches at the (A, B, C, DR, DQ) antigens, *ω*, KDPI, and patient age. We draw 200 pseudo samples, each imputing *ω*conditional on low-resolution (A, B, DR). Table S1 reports the average logit coefficients and the average standard errors across the 200 pseudo samples. Table S2 shows the analogous table when imputing based on low-resolution (A,B, C, DR, DQ).

**Table S1:** Average Logit Coefficients: RMI Mismatches and Five-Year Survival, Imputation conditional on low-resolution (A, B, DR)

|  | Observed Five-Year Survival |  |  | Synthetic Five-Year Survival |  |  |
| --- | --- | --- | --- | --- | --- | --- |
|  | White | Black | Hispanic | White | Black | Hispanic |
| Mismatches 0-10 | -.0043<br>(.0255) | .0193<br>(.0373) | -.0445<br>(.036) | -.0893<br>(.0262) | -.1207<br>(.0381) | -.1422<br>(.0368) |
| KDPI | -.0149<br>(.0024) | -.0108<br>(.0025) | -.012<br>(.0028) | -.0119<br>(.0023) | -.0131<br>(.0024) | -.0069<br>(.0026) |
| Age | -.0171<br>(.005) | .0047<br>(.0048) | -.0071<br>(.0049) | -.0125<br>(.0048) | .003<br>(.0048) | -.011<br>(.0047) |
| Constant | 2.9008<br>(.3335) | 1.3114<br>(.382) | 2.7212<br>(.3514) | 2.9526<br>(.3157) | 2.5718<br>(.3892) | 3.2819<br>(.3789) |
| Observations | 1,496 | 1,370 | 1,416 | 1,496 | 1,370 | 1,416 |

**Table S2:** Average Logit Coefficients: RMI Mismatches and Five-Year Survival, Imputation conditional on low-resolution (A, B, C, DR, DQ)

|  | Observed Five-Year Survival |  |  | Synthetic Five-Year Survival |  |  |
| --- | --- | --- | --- | --- | --- | --- |
|  | White | Black | Hispanic | White | Black | Hispanic |
| Mismatches 0-10 | -.0078<br>(.0261) | .0165<br>(.0378) | -.0514<br>(.0366) | -.1245<br>(.0277) | -.1616<br>(.042) | -.1493<br>(.0377) |
| KDPI | -.0149<br>(.0024) | -.0109<br>(.0025) | -.012<br>(.0028) | -.0147<br>(.0024) | -.0113<br>(.0024) | -.0093<br>(.0026) |
| Age | -.0171<br>(.005) | .0048<br>(.0048) | -.007<br>(.0049) | -.0119<br>(.0047) | .004<br>(.0046) | -.0067<br>(.0044) |
| Constant | 2.9226<br>(.3335) | 1.3344<br>(.3829) | 2.7732<br>(.3606) | 3.3625<br>(.3159) | 2.7181<br>(.4073) | 3.2824<br>(.3689) |
| Observations | 1,496 | 1,370 | 1,416 | 1,496 | 1,370 | 1,416 |

We evaluate imputation of most prevalent types by imputing the most common high resolution antigens for each donor and patient and treating these as data, then estimating the analogous logistic regressions that include age and KDPI. Table S3 shows the logit coefficients of these regressions. Table S4 shows the analogous table when imputing based on low resolution (A, B, C, DR, DQ).

**Table S3:** Logit Coefficients Imputation of Most Prevalent Types for High Resolution Mismatches, Imputation conditional on low-resolution (A, B, DR)

|  | Observed Five-Year Survival |  |  | Synthetic Five-Year Survival |  |  |
| --- | --- | --- | --- | --- | --- | --- |
|  | White | Black | Hispanic | White | Black | Hispanic |
| Mismatches 0-10 | -0.0014<br>(0.0242) | 0.0323<br>(0.0328) | -0.0394<br>(0.0331) | -0.0860<br>(0.0247) | -0.1116<br>(0.0347) | -0.1440<br>(0.0338) |
| KDPI | -0.0150<br>(0.0024) | -0.0108<br>(0.0025) | -0.0120<br>(0.0028) | -0.0118<br>(0.0023) | -0.0131<br>(0.0024) | -0.0068<br>(0.0026) |
| Age | -0.0171<br>(0.0050) | 0.0048<br>(0.0048) | -0.0070<br>(0.0049) | -0.0124<br>(0.0048) | 0.0028<br>(0.0048) | -0.0109<br>(0.0047) |
| Constant | 2.8816<br>(0.3271) | 1.2094<br>(0.3532) | 2.6719<br>(0.3287) | 2.9100<br>(0.3060) | 2.4903<br>(0.3642) | 3.2658<br>(0.3594) |
| Observations | 1,496 | 1,370 | 1,416 | 1,496 | 1,370 | 1,416 |

Robust standard errors in parentheses.

**Table S4:** Logit Coefficients Imputation of Most Prevalent Types for High Resolution Mismatches, Imputation conditional on low-resolution (A, B, C, DR, DQ)

|  | Observed Five-Year Survival |  |  | Synthetic Five-Year Survival |  |  |
| --- | --- | --- | --- | --- | --- | --- |
|  | White | Black | Hispanic | White | Black | Hispanic |
| Mismatches 0-10 | 0.0021<br>(0.0251) | 0.0279<br>(0.0358) | -0.0465<br>(0.0345) | -0.1248<br>(0.0268) | -0.1605<br>(0.0409) | -0.1362<br>(0.0357) |
| KDPI | -0.0150<br>(0.0024) | -0.0109<br>(0.0025) | -0.0120<br>(0.0027) | -0.0147<br>(0.0024) | -0.0113<br>(0.0024) | -0.0093<br>(0.0026) |
| Age | -0.0171<br>(0.0050) | 0.0048<br>(0.0048) | -0.0070<br>(0.0049) | -0.0116<br>(0.0047) | 0.0038<br>(0.0046) | -0.0067<br>(0.0044) |
| Constant | 2.8602<br>(0.3268) | 1.2442<br>(0.3709) | 2.7299<br>(0.3428) | 3.3385<br>(0.3096) | 2.7081<br>(0.3993) | 3.1644<br>(0.3539) |
| Observations | 1,496 | 1,370 | 1,416 | 1,496 | 1,370 | 1,416 |

Robust standard errors in parentheses.

Table S5 is analogous to Table 2 of the main text when conditioning on low resolution (A, B, C, DR, DQ).

**Table S5:** Estimated Effect of an Increase in Mismatches on Survival Probability, Imputation conditional on low-resolution (A, B, C, DR, DQ)

|  | Observed Five-Year Survival |  |  | Synthetic Five-Year Survival |  |  |
| --- | --- | --- | --- | --- | --- | --- |
|  | White | Black | Hispanic | White | Black | Hispanic |
| Most Prevalent Types | 0.0004<br>(0.0044) | 0.0056<br>(0.0079) | -0.0059<br>(0.0033) | -0.0135<br>(0.0013) | -0.0138<br>(0.0007) | -0.0113<br>(0.0008) |
| RMI | -.0013<br>(.0042) | .0033<br>(.0077) | -.0062<br>(.0033) | -.0133<br>(.0013) | -.0137<br>(.0008) | -.0113<br>(.0007) |
| Observations | 1,496 | 1,370 | 1,416 | 1,496 | 1,370 | 1,416 |

Robust standard errors in parentheses.

Table 2 shows the average logit effect for all 200 draws of RMI. Each draw from RMI provides a unique estimate for this effect. Figures S1 and S2 show the complete distributions of estimates resulting from RMI for the effect of increasing mismatches by 1 for an individual with 0 mismatches, KDPI of 50, and age of 50. We show the distributions in figures S1 (with real survival) and S2 (with synthetic survival).

**Figure S1.**
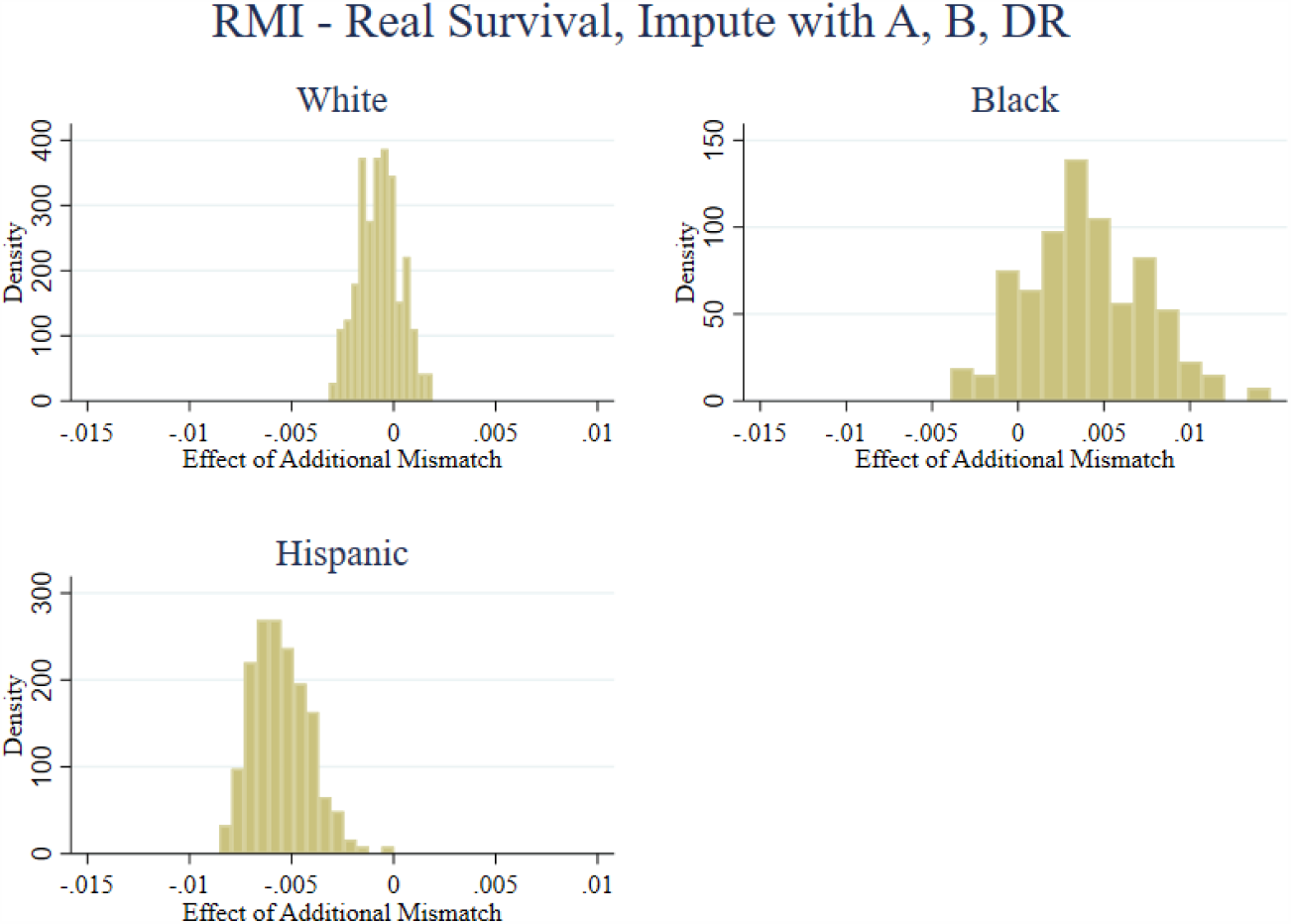

**Figure S2.**
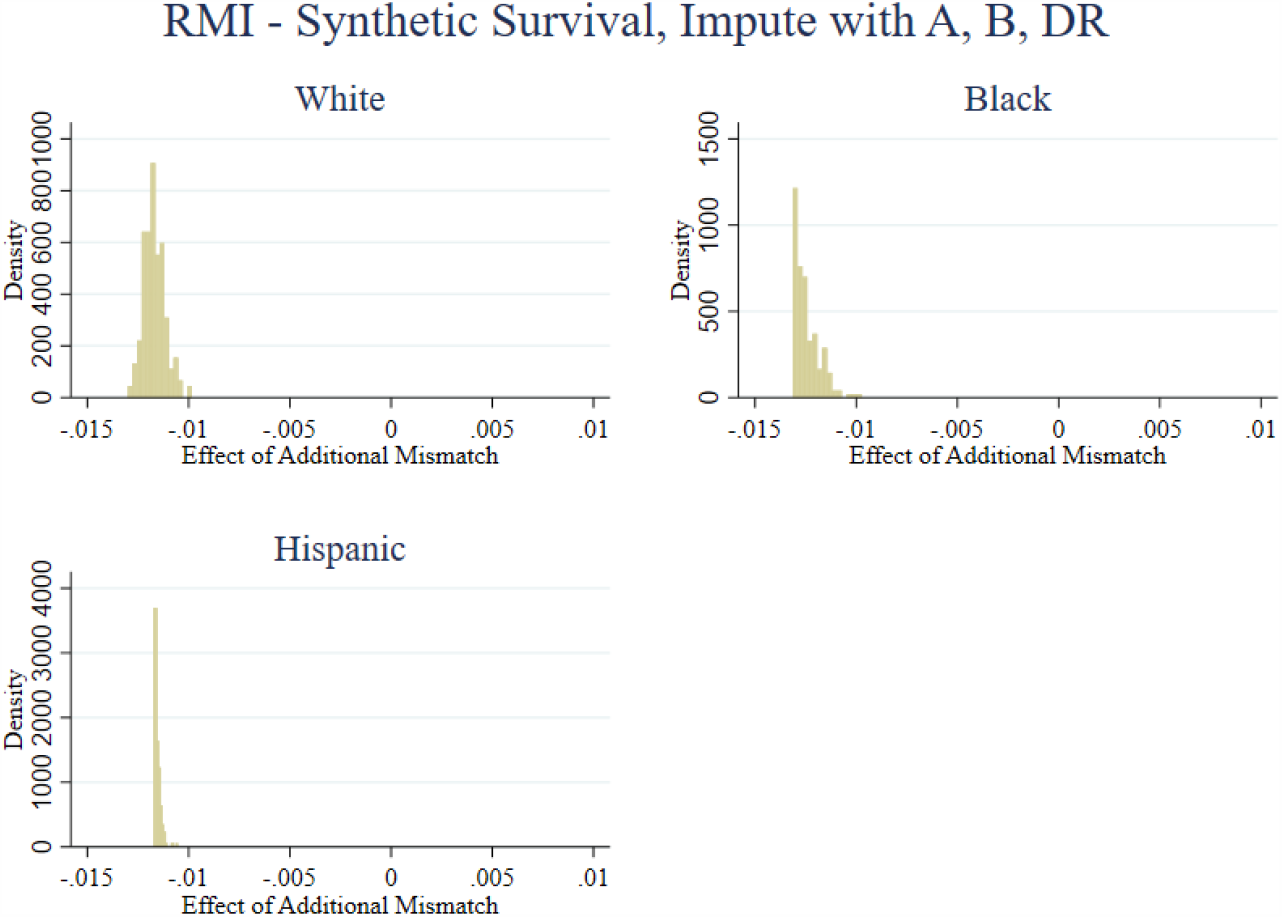

Figures S3 and S4 show effects when imputing is based on low resolution (A, B, DR, C, DQ). S3 uses real survival and S4 uses synthetic survival.

**Figure S3.**
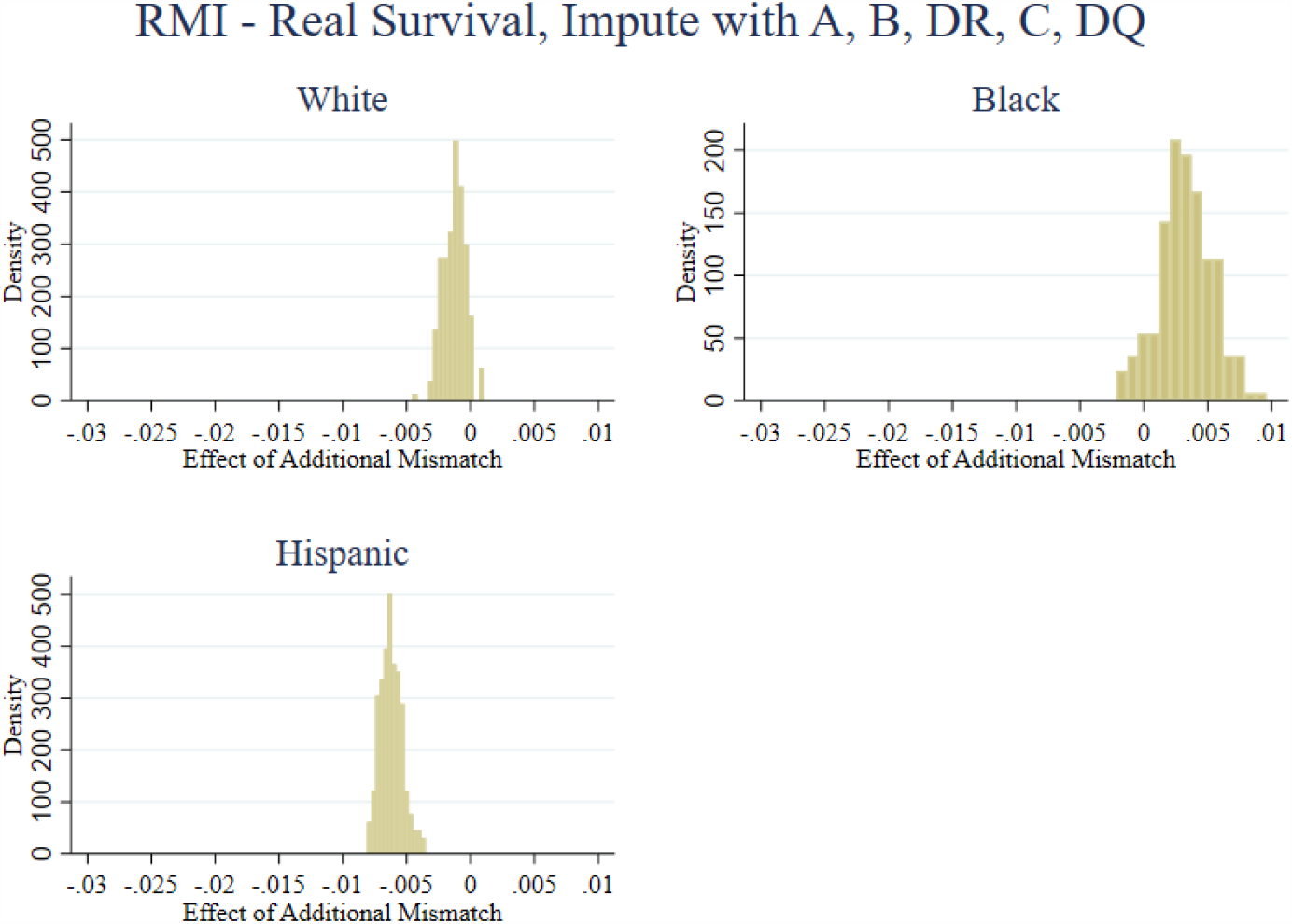

**Figure S4.**
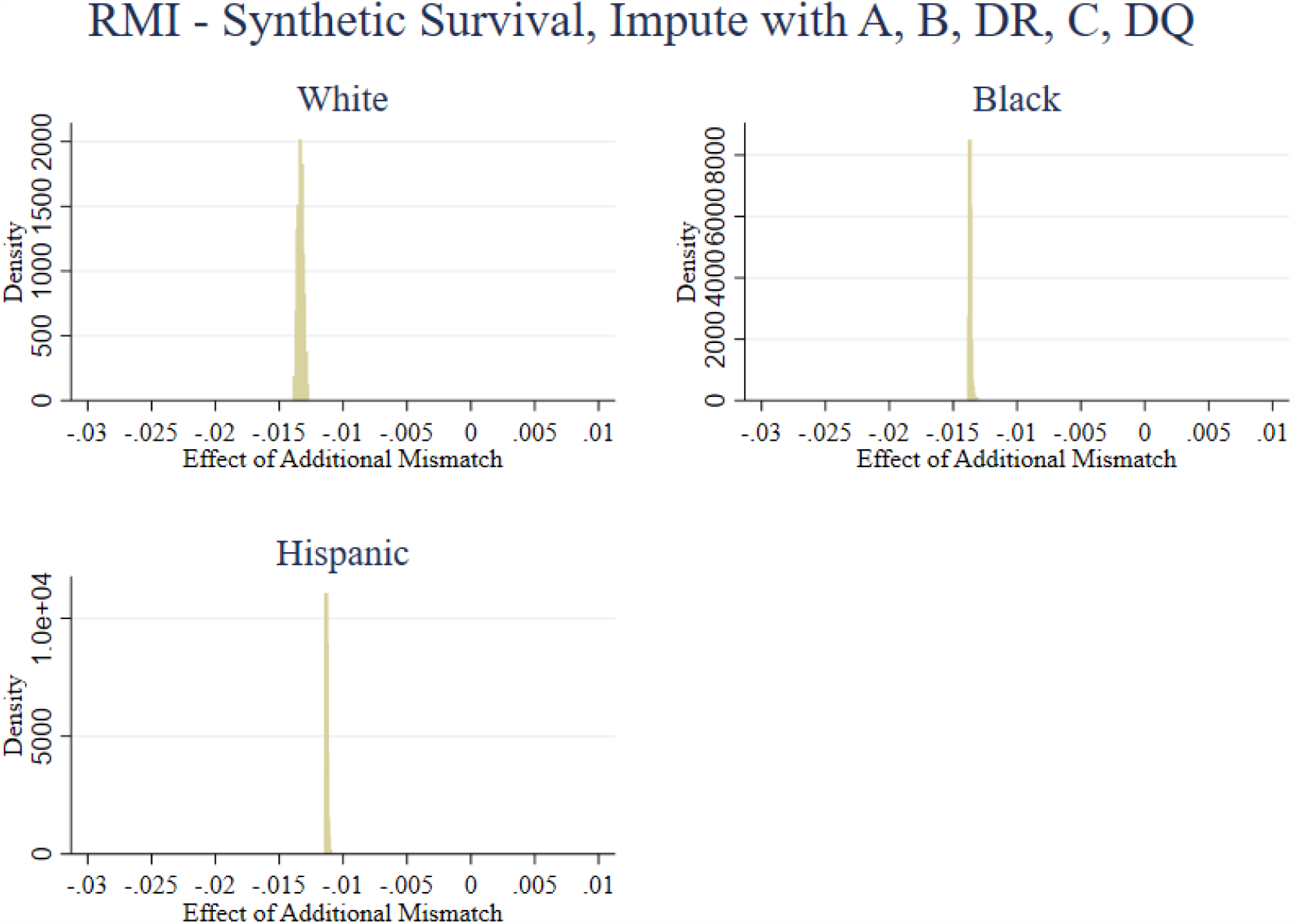

### Supplementary Material 2 – Imputing the Expected Number of Mismatches

We estimate logit models predicting survival as a function of the expected number of four-digit mismatches, age of the patient, and KDPI. Expected values are calculated based on the low-resolution (A, B, DR) antigens for patients and donors. Table S6 shows the logit coefficients. Table S7 is analogous to Table S6 when calculating expected mismatches conditional on low resolution (A, B, C, DR, DQ). Results are qualitatively similar to other analysis regarding RMI and imputation of most prevalent types.

**Table S6:**
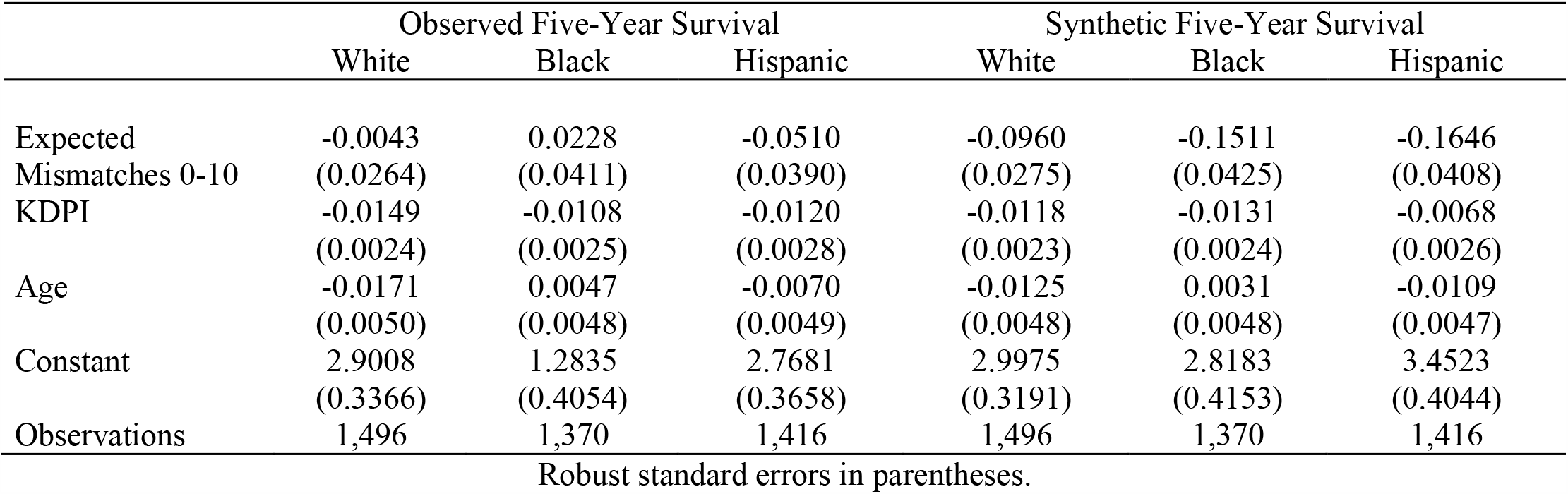
Logit Coefficients: Expected Mismatches and Five-Year Survival, Imputation conditional on low-resolution (A, B, DR)

**Table S7:**
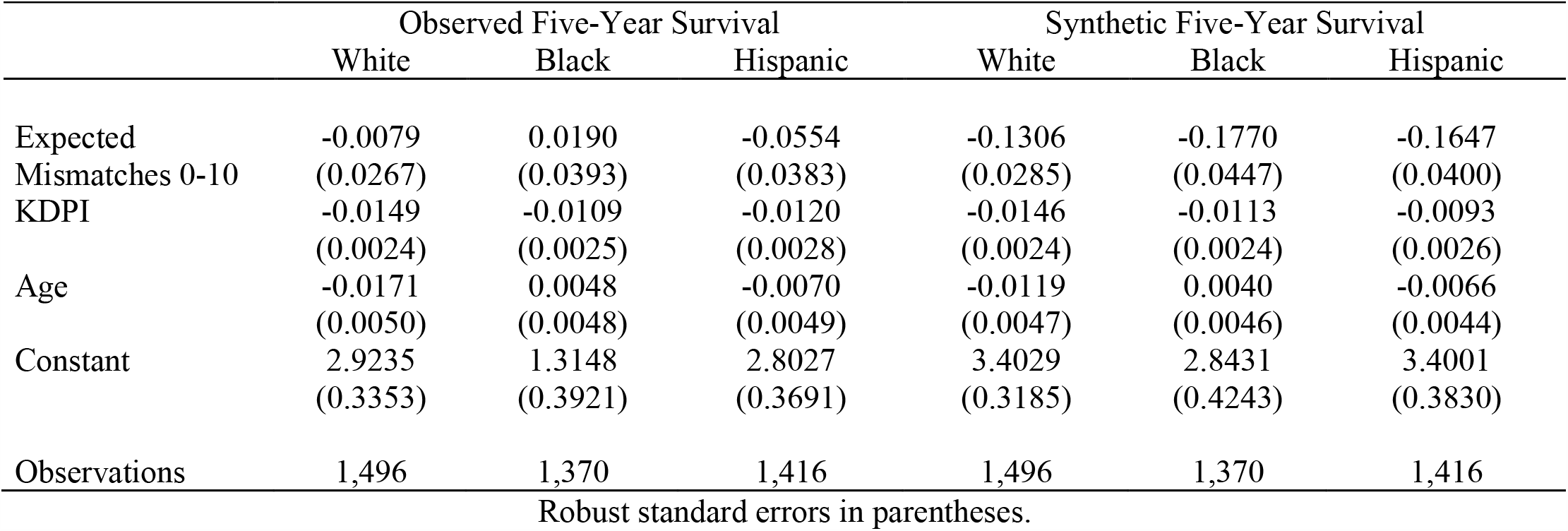
Logit Coefficients: Expected Mismatches and Five-Year Survival, Imputation conditional on low-resolution (A, B, C, DR, DQ)

### Supplementary Material 3 – Linear Regression Results

The results throughout the main text and other supplementary materials use logistic regression. This section shows analogous results using linear regression. Tables S8, S9, S10, and S11 are respectively analogous to tables 1, S1, S3, and S6. Because linear models have coefficients equivalent to marginal effects, there is no presentation of an analogy to table 2.

**Table S8:** Observed Low Resolution Mismatches and Five-Year Survival

|  | Observed Five-Year Survival |  |  | Synthetic Five-Year Survival |  |  |
| --- | --- | --- | --- | --- | --- | --- |
|  | White | Black | Hispanic | White | Black | Hispanic |
| Mismatches 0-10 | -0.0200<br>(0.0040) | -0.0205<br>(0.0060) | -0.0170<br>(0.0040) | -0.0200<br>(0.0040) | -0.0205<br>(0.0060) | -0.0170<br>(0.0040) |
| KDPI | -0.0027<br>(0.0004) | -0.0021<br>(0.0005) | -0.0015<br>(0.0004) | -0.0027<br>(0.0004) | -0.0021<br>(0.0005) | -0.0015<br>(0.0004) |
| Age | -0.0019<br>(0.0007) | 0.0007<br>(0.0008) | -0.0010<br>(0.0006) | -0.0019<br>(0.0007) | 0.0007<br>(0.0008) | -0.0010<br>(0.0006) |
| Observations | 1,496 | 1,370 | 1,416 | 1,496 | 1,370 | 1,416 |

Robust standard errors in parentheses.

**Table S9:** Average Regression Coefficients: RMI Mismatches and Five-Year Survival

|  | Observed Five-Year Survival |  |  | Synthetic Five-Year Survival |  |  |
| --- | --- | --- | --- | --- | --- | --- |
|  | White | Black | Hispanic | White | Black | Hispanic |
| Mismatches 0-10 | -.0012<br>(.0041) | .0035<br>(.0066) | -.0063<br>(.0046) | -.0152<br>(.0041) | -.0202<br>(.0059) | -.0201<br>(.0045) |
| KDPI | -.0026<br>(.0004) | -.0019<br>(.0004) | -.0019<br>(.0004) | -.0022<br>(.0004) | -.0024<br>(.0004) | -.0012<br>(.0004) |
| Age | -.0025<br>(.0007) | .0008<br>(.0008) | -.001<br>(.0006) | -.0021<br>(.0007) | .0005<br>(.0008) | -.0017<br>(.0007) |
| Observations | 1,496 | 1,370 | 1,416 | 1,496 | 1,370 | 1,416 |

**Table S10:**
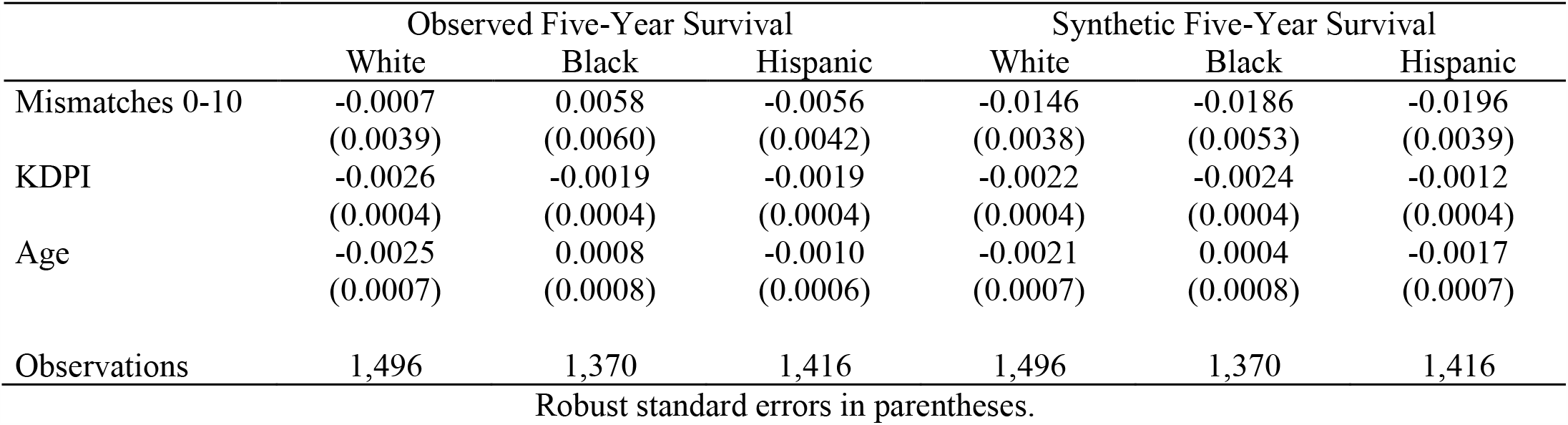
Imputation of Most Prevalent Types, High Resolution Mismatches and Survival.

|  | Observed Five-Year Survival |  |  | Synthetic Five-Year Survival |  |  |
| --- | --- | --- | --- | --- | --- | --- |
|  | White | Black | Hispanic | White | Black | Hispanic |
| Mismatches 0-10 | -0.0007<br>(0.0039) | 0.0058<br>(0.0060) | -0.0056<br>(0.0042) | -0.0146<br>(0.0038) | -0.0186<br>(0.0053) | -0.0196<br>(0.0039) |
| KDPI | -0.0026<br>(0.0004) | -0.0019<br>(0.0004) | -0.0019<br>(0.0004) | -0.0022<br>(0.0004) | -0.0024<br>(0.0004) | -0.0012<br>(0.0004) |
| Age | -0.0025<br>(0.0007) | 0.0008<br>(0.0008) | -0.0010<br>(0.0006) | -0.0021<br>(0.0007) | 0.0004<br>(0.0008) | -0.0017<br>(0.0007) |
| Observations | 1,496 | 1,370 | 1,416 | 1,496 | 1,370 | 1,416 |

Robust standard errors in parentheses.

**Table S11:** Expected Mismatches and Five-Year Survival

|  | Observed Five-Year Survival |  |  | Synthetic Five-Year Survival |  |  |
| --- | --- | --- | --- | --- | --- | --- |
|  | White | Black | Hispanic | White | Black | Hispanic |
| Expected Mismatches 0-10 | -0.0012<br>(0.0043) | 0.0040<br>(0.0074) | -0.0072<br>(0.0049) | -0.0163<br>(0.0042) | -0.0248<br>(0.0063) | -0.0227<br>(0.0048) |
| KDPI | -0.0026<br>(0.0004) | -0.0019<br>(0.0004) | -0.0019<br>(0.0004) | -0.0022<br>(0.0004) | -0.0024<br>(0.0004) | -0.0012<br>(0.0004) |
| Age | -0.0025<br>(0.0007) | 0.0008<br>(0.0008) | -0.0010<br>(0.0006) | -0.0021<br>(0.0007) | 0.0005<br>(0.0008) | -0.0017<br>(0.0007) |
| Observations | 1,496 | 1,370 | 1,416 | 1,496 | 1,370 | 1,416 |

Robust standard errors in parentheses.

## Abbreviations

HLA: Human Leukocyte Antigen
KDPI: Kidney Donor Profile Index
OPTN: Organ Procurement and Transplantation Network
RMI: Random Multiple Imputation
SRTR: Scientific Registry of Transplant Recipients

## Notes

### Competing Interest Statement

The authors have declared no competing interest.

### Funding Statement

No external funding was received.

### Author Declarations

This study was reviewed and exempted from approval by the Northwestern University Institutional Review Board.

